# Who funds stroke trials in Europe? A survey of funding sources for randomised controlled stroke trials by the European Stroke Organisation Trials Alliance (ESOTA) network

**DOI:** 10.64898/2026.06.22.26356203

**Authors:** Smriti Agarwal, Jean Marc Olivot, Christine Roffe, Michael Knoflach

## Abstract

**Aims and scope:** Evidence from randomised controlled trials (RCTs) has transformed stroke care. There are no systematically collected data on the amount of public funding, critical to delivering trials, going into stroke RCTs. To understand the extent of stroke RCT funding by national and EU funding bodies across Europe, the European Stroke Organisation Trials Alliance (ESOTA) conducted a survey of its member nations.

**Methods:** This is an observational study of research funding in Europe. The ESOTA steering group sent an electronic survey to the leads of the 16 participating national networks from 14 countries. Structured survey questions included who the funding bodies were in each country, the number of RCT applications put forward for public national or EU funding, the number of successful and failed applications, and the amount of funding granted between 01/01/2022 and 31/12/2023.

**Results:** Responses were received from 13 of 14 participating countries. There was significant variation in the number of grant applications submitted by individual countries, ranging from 0-17 during the 24-month survey period. The median number of funded studies per country was 1 (IQR 3, range 0-9) representing a median success rate of 47.1 % (IQR 21.1-59.4%), with no RCTs granted joint European funding.

**Conclusions:** Our survey highlights significant inequities in stroke trial funding across Europe. Given the encouraging rate of successful applications overall, it is important for all member networks to submit proposals. This is particularly pertinent for multicentre trials, given the evolution of evidence base in stroke towards large trials, across diverse populations.

## Introduction

Evidence from randomised controlled trials (RCTs) has driven significant improvements in stroke care over the last 2 decades^1^, with thrombolysis, thrombectomy and stroke unit care being only a few examples. Stroke is a leading cause of adult disability with 12.2 million new strokes every year^2^ and 9.5 million stroke survivors across Europe^3^. Public funding for RCTs has historically been insufficient in general^4^, and particularly, in stroke^5^. Documenting gaps in funding available and strategies to address these gaps are critical to maintaining scientific rigour and transparency that may uniquely come from non-commercially funded RCTs^6,7^. Investment of public money into RCTs is cost-effective for health systems by reducing overall health costs and improving health indices at the population level^8,9^.

The European Stroke Organisation Trials Alliance (ESOTA https://eso-trialsalliance.org/) was set up in 2019 as a Europewide network, under the umbrella of the European Stroke Organisation (ESO), comprising of national network leads alongside a steering group. The key aim of the ESOTA is to provide a sustainable, long-term, collaborative infrastructure to support investigator-led stroke randomised trials in Europe. So far, no clear data exists on the amount of funding going into stroke RCTs across Europe. In particular, there is no reliable data on investigator-led stroke RCTs with public funding. The current study was conducted as a structured survey by the ESOTA to document the landscape of RCT funding across European trial networks and assess the need for specific funding bodies to support trial activity. This data would be key in the efforts of the ESO to promote stroke trials to funders and foster collaboration across Europe.

## Methods

In order to systematically evaluate the landscape of stroke RCT funding by national and EU funding bodies across Europe, the ESOTA designed an electronic survey https://survey.lamapoll.de/ESO_ESOTA_Survey-project_2024 that was completed by leads of individual member networks. The responses from the Austrian network were included, where membership procedures are ongoing. The time-period covered by the survey was 1^st^ January 2022 -31^st^ December 2023.

The following items were addressed in the survey:

1. The number of RCT applications made during the survey period
2. The number or RCTs which received funding approval during the survey period

The questions included in the survey are listed below, all referring to the 24-month time period (1^st^ January 2022 -31^st^ December 2023).

- Were stroke RCTs applied for public funding in the network country?
- How many stroke RCTs were funded in the country with national or EU public funding?
- Name of the top 3 relevant funding agencies for RCTs in the network country
- How would you rate the possibilities for funding stroke RCTs in your network country (Likert scale 1 good – 5 bad)?

In addition, the ESOTA also provided a template letter (with appropriately translated versions) to the national leads, who were asked to approach their country’s key funding bodies to obtain this information (supplement for letter template). We highlight data from this part of the study from the UK, where detailed responses were received from three funding bodies. The time-period covered varied slightly compared with the main survey (2021-2022 and 2022-2023) due to logistics of the relevant funding cycles. The questions for the funding body survey are listed below.

- How many stroke RCT applications did you receive in this period?
- How many stroke RCT applications were successfully funded in this period?
- List of successfully funded RCTs in this period
- Have you funded any RCTs jointly with European funding in this period?
- How many RCT applications overall did you receive in this period?
- How many of these were successfully funded?

The success rate of applications was also examined against the individual network country’s gross domestic product (GDP) and incidence of stroke. The data for the latter was derived from https://www.statista.com/.

Descriptive statistics are presented for the survey findings. Success rates were calculated by dividing the number of applications submitted by the number of funded applications and expressed in %. The median success rate of all countries was calculated from those countries with at least one RCT submitted for funding.

## Results

Survey responses were received for 13 out of 14 European countries whose national trial leads were approached (Germany, Belgium, Greece, Switzerland, Austria, France, Ireland, United Kingdom (England/Wales and Scotland) Netherlands, Turkey, Italy, Spain, Estonia and Czech Republic) as shown in table 1. The network structures in countries varies slightly with 2 networks in Greece and 2 in the UK.

**Table 1.**
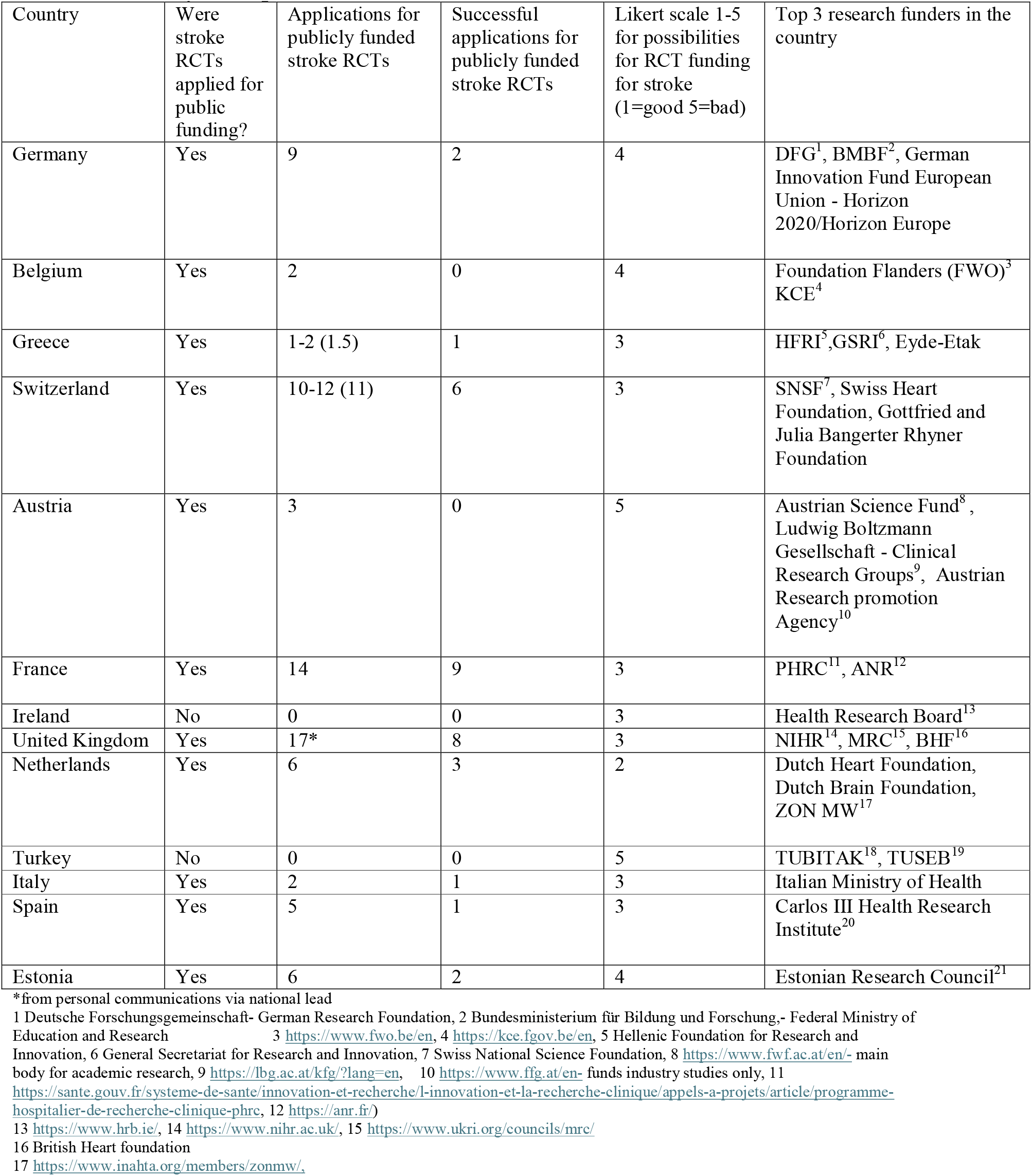
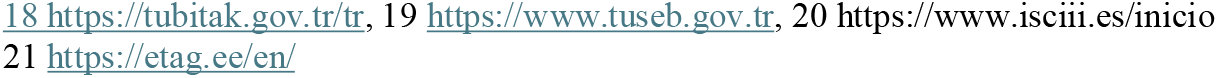
Summary of responses received from national clinical trial network leads.

| Country | Were stroke RCTs applied for public funding? | Applications for publicly funded stroke RCTs | Successful applications for publicly funded stroke RCTs | Likert scale 1-5 for possibilities for RCT funding for stroke (1=good 5=bad) | Top 3 research funders in the country |
| --- | --- | --- | --- | --- | --- |
| Germany | Yes | 9 | 2 | 4 | DFG<sup>1</sup>, BMBF<sup>2</sup>, German Innovation Fund European Union - Horizon 2020/Horizon Europe |
| Belgium | Yes | 2 | 0 | 4 | Foundation Flanders (FWO)<sup>3</sup> KCE<sup>4</sup> |
| Greece | Yes | 1-2 (1.5) | 1 | 3 | HFRI<sup>5</sup>, GSRI<sup>6</sup>, Eyde-Etak |
| Switzerland | Yes | 10-12 (11) | 6 | 3 | SNSF<sup>7</sup>, Swiss Heart Foundation, Gottfried and Julia Bangerter Rhyner Foundation |
| Austria | Yes | 3 | 0 | 5 | Austrian Science Fund<sup>8</sup>, Ludwig Boltzmann Gesellschaft - Clinical Research Groups<sup>9</sup>, Austrian Research promotion Agency<sup>10</sup> |
| France | Yes | 14 | 9 | 3 | PHRC<sup>11</sup>, ANR<sup>12</sup> |
| Ireland | No | 0 | 0 | 3 | Health Research Board<sup>13</sup> |
| United Kingdom | Yes | 17* | 8 | 3 | NIHR<sup>14</sup>, MRC<sup>15</sup>, BHF<sup>16</sup> |
| Netherlands | Yes | 6 | 3 | 2 | Dutch Heart Foundation, Dutch Brain Foundation, ZON MW<sup>17</sup> |
| Turkey | No | 0 | 0 | 5 | TUBITAK<sup>18</sup>, TUSEB<sup>19</sup> |
| Italy | Yes | 2 | 1 | 3 | Italian Ministry of Health |
| Spain | Yes | 5 | 1 | 3 | Carlos III Health Research Institute<sup>20</sup> |
| Estonia | Yes | 6 | 2 | 4 | Estonian Research Council<sup>21</sup> |

\*from personal communications via national lead
1 Deutsche Forschungsgemeinschaft- German Research Foundation, 2 Bundesministerium für Bildung und Forschung,- Federal Ministry of Education and Research
3 https://www.fwo.be/en, 4 https://kce.fgov.be/en, 5 Hellenic Foundation for Research and Innovation, 6 General Secretariat for Research and Innovation, 7 Swiss National Science Foundation, 8 https://www.fwf.ac.at/en/-main
body for academic research, 9 https://lbg.ac.at/kfg/?lang=en, 10 https://www.ffg.at/en- funds industry studies only, 11 https://sante.gouv.fr/systeme-de-sante/innovation-et-recherche/l-innovation-et-la-recherche-clinique/appels-a-projets/article/programme-hospitalier-de-recherche-clinique-phrc, 12 https://anr.fr/
https://www.hrb.ie/, 14 https://www.nihr.ac.uk/, 15 https://www.ukri.org/councils/mrc/
British Heart foundation
https://www.inahta.org/members/zonmw/.
https://tubitak.gov.tr/tr, 19 https://www.tuseb.gov.tr, 20 https://www.isciii.es/inicio https://etag.ee/en/

The number of applications varied from 0-17 with a mean of 5.8 (SD 5.4) and median of 4.5 (IQR 2-9). The distribution of funding applications was uneven across countries, with no RCT applications made in 2 networks (Ireland and Turkey) over the 24-month study period and more than 10 in 3 countries (United Kingdom, France and Switzerland). Success rate of applications varied from 0-66.7% in the networks where applications were made for investigator led RCT funding from public bodies. The mean Likert scale reading of the perception of possibilities of funding in the individual network countries ranged from 2-5 with a mean of 3.5 (SD 0.9) and a median of 3. There was a trend towards a higher success rate in countries with higher GDP (figure 1a) and lower rate amongst those with higher stroke incidence (figure 1b). Quantitative statistics were not performed here given the small numbers. No EU cross-border co-funded RCTs were identified from the survey

**Figure 1.**
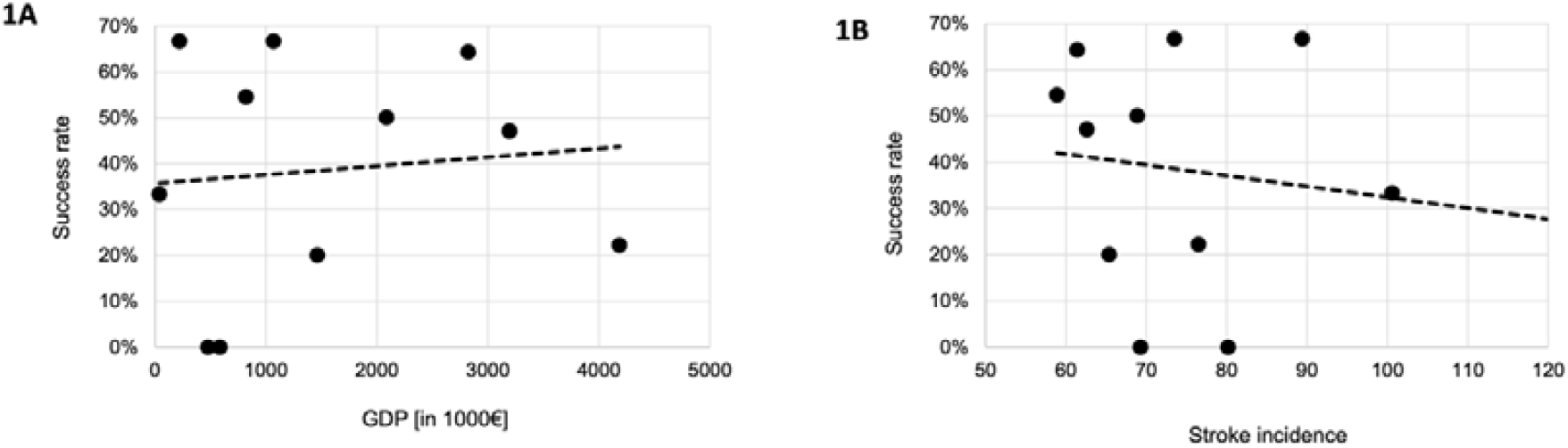
RCT application success rate in context of national gross domestic product (GDP) and stroke incidence. Figure 1a % of successful stroke randomized control trial (RCT) applications plotted against the individual network country’s GDP (million Euros, source Statista.com) Figure 1b % of successful stroke randomized control trial (RCT) applications plotted against stroke incidence per 100,000 population (source Statista.com) in each individual network country

Responses from individual funding bodies were only available for the UK trials network (Table 2). 5 funding bodies relevant for stroke research (National Institute for Health and Care Research-NIHR, Medical Research Council -MRC, Wellcome Trust-WT, British Heart Foundation-BHF, Stroke Association-SA, and Chest Heart Stroke Scotland-CHSS) were approached individually for detailed information regarding stroke RCT funding in the survey time period. The BHF, WT and CHSS provided detailed responses. The SA did not fund RCTs, but provided details of other stroke studies and career development programmes for stroke researchers. The NIHR provided a link to their database of funded RCTs, but was not able to provide information on number of trial applications received in stroke and applications received overall. An estimate of submitted applications was made by personal communication by the UK national lead.

**Table 2.** UK funding bodies and stroke RCT funding.

|  | Overall RCT applications received | Stroke RCT applications received | Overall RCTs Successfully funded | Stroke RCTs Successfully funded | Joint stroke RCTs with European funding |
| --- | --- | --- | --- | --- | --- |
| **2021/2022** |  |  |  |  |  |
| Wellcome Trust | 88 | 1 | 20 | 0 | 0 |
| British Heart Foundation | 9 | 1 | 5 | 0 | 0 |
| Medical Research Council | * | * | * | 0 | 0 |
| **2022/2023** |  |  |  |  |  |
| Wellcome Trust | 84 | 0 | 37 | 0 | 0 |
| British Heart Foundation | 4 | 1 | 2 | 1 | 0 |
| Medical Research Council | * | * | * | 0 | 0 |

\*information unavailable

The overall success rate across the three funding bodies which received stroke RCT applications was 33.1% in the survey period. The BHF received 2 stroke RCT applications, of which 1 was funded. The Wellcome trust received 1 stroke RCT application in the two-year period (out of 172, overall RCT applications) and it was not funded. The MRC did not fund any RCTs during this period. While the NIHR funded 4 stroke RCTs during this period, information about overall applications received and success rates of stroke RCTs, per se, was not available directly. Overall, approximately 0.5% of RCT applications received by the Wellcome Trust and 15% of those received overall by the BHF were stroke RCTs. None were funded by the WT and 14% of successfully funded RCTs by the BHF were in stroke.

There were no joint European or cross-border funding supported trials in the time-period covered by the survey.

## Discussion

Our study highlights the unmet need for stroke research and major inequities in stroke trial funding across Europe. Public funding for RCTs does not reflect its incidence and varies significantly between countries, with two network countries not making any RCT applications for public funding and a further 2 having no publicly funded stroke trials, in the two-year period covered by the survey. This is particularly pertinent for multicentre trials, given the evolution of evidence base in stroke towards large trials across diverse populations.

The variability across networks in the number of applications made for support from public bodies, as well as success rates, highlights the need for closer collaboration between network member countries. Additionally, stroke funding opportunities sit on generic, nation-specific research funding infrastructure, which probably accounts for a significant part of the variation. The data on stroke incidence and RCT funding success highlights the need for trials in populations that have higher disease burden. Networks like the ESOTA could provide support to reduce inequities in RCT funding. Specifically, the ESOTA could be a forum for pre-screening trial applications and contribute to an informal peer review process, especially to encourage less experienced investigators. It could also host a data hub for aggregated grant applications and outcomes.

The more detailed survey of UK funders revealed even larger gaps with the charity funders we approached. This information was not available for other individual funders in other network countries, so we are unable to investigate whether these gaps are seen more widely. This is one limitation of the study, which also highlights the fact that transparent information about funding support for RCTs is difficult to find. Organisations such as the Dutch Heart Foundation have started a specific programme for stroke RCTs since 2021 and are willing to support national arms of multinational trials, alongside national trials. There is a need for wider availability of such information to enable success of clinical trials.

While the success rates from the survey may appear encouraging overall, there are some caveats to the interpretation of the results. Number of applications made are likely to have been underestimated due to a degree of recall bias. Also, a strict definition of what would be classified as a stroke RCT is difficult, given the nature of the field, and there would be anticipated overlaps with cardiovascular trials. Hence, our success rates should be considered as maximum estimates rather than necessarily a complete reflection of the funding scenario.

Poor access to funding is a major barrier to bringing evidence-based effective interventions to patients^10^ and could deter investigators, given high costs of research from the planning stage to execution^11^. Improving access to public funding is an important aspirational goal highlighted by national bodies to improve research and outcomes for patients^12^.

Moreover, stroke funding lags behind other conditions, such as cancer. Just under a third (29.4 %) of all clinical trial submissions to the UK Medicines and Healthcare products Regulatory Agency were in cancer, compared with 12.2% in neurological disorders overall and 5.2% in cardiovascular diseases according to a recent study, with no specific data available for stroke trials per se. ^10^ Our study results from the UK funding bodies supports this, though we do not have information on specific comparisons with other speciality areas. There is a need for organisations such as the ESO to campaign for funding into stroke RCTs.

Our study has some limitations. The data was obtained from network leads rather than publicly available databases. This could introduce a degree of variability in the data collection methodology. There could be a degree of self-reporting due to the inherent nature of the survey. This is also an issue with the interpretation of success rates for the UK funding body data, given some of the information was obtained by personal communication by the national lead, and hence likely to underestimate of the actual number of applications made. However, this also highlights the need for an open access repository of public funded resources supporting stroke RCTs which will improve transparency and help investigators target relevant funders.

Our work highlights the need for transparency in resource allocation to RCT’s in Europe and more widely. One potential solution could be a repository where RCT applications made with funding details could be, prospectively, recorded. Efforts could focus on encouraging funders to transparently and prospectively report applications received and funded on an annual basis, into such a repository. Networks, such as the ESOTA, with wider partners such as the Global Alliance of Independent Networks focused on Stroke trials (GAINS), would be crucial in informing policy in this area, by aligning metrics and helping to standardise calls.

Engagement in clinical trials has been shown to improve patient outcomes.^13,14^ Stroke trials will bring results needed to shape practice, inform guidelines^15^, improve stroke services and outcomes. Research needs to become part of routine clinical practice, rather than just being an add-on for specialist and academic teams. To achieve this, we need more stroke neurologists becoming researchers, more grant applications, but also a higher proportion of research funding going to stroke studies. ESOTA can provide researchers with resources, guidance and contacts for national and international collaborations, in a model similar to the Global Cardiovascular Research Funders Forum (GCRFF)^16^ with a wider Europewide reach and a specific focus on stroke.

## Conclusions

There are significant inequities in public funding into stroke RCTs which are a potential barrier to bringing effective interventions to patients. With large variations in applications made alongside reasonable success rates overall, there is a need for networks to collaborate and submit proposals. Networks such as the ESOTA, working alongside wider global partner networks, have a role to campaign for promoting wider collaboration, supporting all networks with funding applications. This will ultimately improve patient outcomes in stroke.

## Supporting information

supplement- details of ESOTA steering and advocacy groups

## Data Availability

All data produced in the present study are available upon reasonable request to the authors

## Notes

### Competing Interest Statement

The authors have declared no competing interest.

