## supplement- details of ESOTA steering and advocacy groups for "Who funds stroke trials in Europe? A survey of funding sources for randomised controlled stroke trials by the European Stroke Organisation Trials Alliance (ESOTA) network"

Supplemental file

Submission on behalf of the

ESOTA steering group, national stroke trial network leads and the ESO advocacy group^**^

**(Peter Kelly^5^, Simona Sacco^6^, Noémie Le Quément^7^, Marcel Arnold^8^, Ethem Murat Arsava^9^, Charlotte Cordonnier^10^, Milani Deb-Chatterji^11^, Maria Alonso De Leciñana Cases^12^, Diederik Dippel^13^, María Mar Freijo^14^, Annette Fromm^15^, Janika Kõrv^16^, Christine Kremer^17^, Lukas Mayer-Sue^4^, Robin Lemmens^18^, Thomas Meinel^19^, Robert Mikulik^20^, Paul Nederkoorn^21^, Christopher Price^22^, Rossana Tassi^23^, Götz Thomalla^24^, Danilo Toni^25^, Georgios Tsivgoulis^26^, Silke Walter^27^, William Whiteley^28^, Andrea Zini^29^ Michèle Schaub Jackson^7^, Guillaume Turc^30^, Barbara Casolla^31^, Hanne Krarup Christensen^32^, Francesca Romana Pezzella^33^, Nicoletta Brunelli^34^, Bart van der Worp^35^

*^5^University College Dublin, Ireland/European Stroke Organisation(ESO), ^6^University of L’Aquila, Italy/ESO, ^7^ESO, ^8^University Hospital Bern (Inselspital), S*witzerl*and, ^9^Hacettepe University, Ankara, Turkey, ^10^University of Lille, INSERM, CHU Lille, U1172- Lille Neuroscience and Cognition, France,* *^11^Dept. Of Neurology, University Hospital Schleswig Holstein Campus Kiel, Kiel, Germany, ^12^La Paz Institute for Health Research (idiPAZ), (La Paz University Hospital- Universidad Autónoma de Madrid) Madrid, Spain, ^13^Erasmus MC, University Medical Center Rotterdam, The Netherlands, ^14^Biobizkaia Health Research Institute. Stroke Unit Cruces University Hospital, Osakidetza, Spain, ^15^Bergen Stroke Research Group*

*Department of Neurology, Haukeland University Hospital, Bergen,* *Norway,^16^ Department of Neurology and Neurosurgery, University of Tartu, Estonia, ^17^Skåne University Hospital Malmö, Department of Clinical Sciences Lund University, Sweden, ^18^KU Leuven & University Hospitals Leuven, Belgium, ^19^Stroke University Hospital Bern (Inselspital), Switzerland, ^20^Health Management Institute, Brno and Tomas Bata Hospital, Zlin, Czech Republic, ^21^Amsterdam University Medical Centers, The Netherlands, ^22^Newcastle University, UK , ^23^* *Stroke Unit, AOU Senese, Italy, ^24^University Medical Center Hamburg-Eppendorf, Germany, ^25^University of Rome, Italy, ^26^National and Kapodistrian University of Athens, Greece, ^27^Saarland University, Germany, ^28^University of Edinburgh, UK, ^29^University of Bologna, Italy, ^30^Sainte-Anne Hospital, Paris, France, ^31^Université Côte d'Azur UR2CA-URRIS, Stroke-Unit, CHU Hôpital Pasteur 2, Nice, France,*

*^32^* *Bispebjerg and Frederiksberg Hospital, University of Copenhagen, Denmark, ^33^Stroke UNit, Department of Neuroscience, San Camillo Forlanini Hospital, Rome, Italy*, *^34^ Vall d' Hebron University Hospital, Barcelona, Spain, ^35^*[*University Medical Center (UMC) Utrecht*](https://researchinformation.umcutrecht.nl/en/organisations/university-medical-center-umc-utrecht/)*, the Netherlands.*
